# Standardization of a High-Fidelity Postpartum Hemorrhage Simulation Scenario Across Two Geographically Separated Campuses

**DOI:** 10.1101/2023.01.31.23285293

**Authors:** Miriam S. Khalepai, Shannon Sturgeon, Andrea Done, Tyler Yates, Mark Wardle, Isain Zapata, Susan Carter

## Abstract

**Introduction:** The utilization of high fidelity simulations across medical school campuses has grown in recent years as simulation is a useful tool in allowing students to practice real life scenarios in a safe space, as well as providing exposure to rarer cases in medicine. This study evaluated second and third-year medical students’ recognition and management of a postpartum hemorrhage (PPH) due to a retained placenta in SimMom®. The objective was to describe the methods and procedures used in PPH simulation to educate these students across two geographically separated medical school campuses. It became apparent that there is a need to standardize medical simulation to maintain the quality of medical education.

**Methods:** A case-scenario simulation with retained tissue had a run-time of 10 minutes. Times were marked when students identified the retained placental tissue was the cause of hemorrhage and when students extracted the fragment. A debrief with faculty followed the scenario regarding performance and education about the “4 T’s”of post-partum hemorrhage differential diagnosis (Tone, Trauma, Tissue, Thrombin), and the students’ feelings towards the simulation were evaluated with a post simulation survey.

**Results:** Despite efforts to maintain consistent simulations between the two campuses, there was a significant difference of timing between the two campuses to successful completion of the simulation, as well as in the number of students that did not successfully complete the goals of the simulation. However, the post-simulation survey did show that students on both campuses found value in participating in the simulation.

**Discussion:** The performance across the two campuses was statistically different, which illustrates a need to standardize simulation education experiences across medical school campuses. However, this protocol was an effective way to educate students and allow students to have hands-on experience in diagnosing and treating a rare but deadly complication of childbirth. This initial study may facilitate expansion on this particular, topic, as well as suggest more simulation education delivery within the medical school curriculum.

## INTRODUCTION

The World Health Organization (WHO) reported that 810 women died from complications related to pregnancy and childbirth daily in 2017. ^1^The leading cause of maternal mortality is postpartum hemorrhage (PPH), ^2^ defined by the American College of Obstetrics and Gynecology (ACOG) as cumulative blood loss of 1,000 mL or greater with symptoms of hypovolemia within 24 hours post-parturition.^3^ Uterine atony is the most common cause of postpartum hemorrhage.^4^ A less-frequent etiology of PPH is retained tissue, such as the placenta or placental fragments, which presents in less than 3% of vaginal deliveries. ^5^ Retained placenta is low on the differential diagnosis as the cause of PPH, and it may be difficult to recognize this etiology in time to prevent significant morbidity and mortality. Laerdal Medical™ has developed the SimMom® manikin, which includes a feature for a retained placental fragment, allowing students to have exposure to an infrequent obstetric complication. A study performed by Deering et al. has shown that students who performed better on simulated deliveries participated in more deliveries during their subsequent OB/GYN rotation.^6^ This study describes the methods and procedures used in a simulation to educate second- and third-year medical students across two geographically separated medical school campuses by timing the students’ recognition and management of a PPH due to a retained placenta; thus, this scenario yields hands-on experience to enhance the confidence of medical students in a rarer part of medicine. ^7^ Moreover, there are 31 medical schools, both allopathic (26)^8^ and osteopathic (5)^9^, that have secondary campuses to accommodate the growing number of medical students. The need to standardize medical simulation experiences across these campuses is important to maintain the quality of medical education.

## METHODS

The SimMom® Gen 2 manikin with Automated Delivery Module (ADM) with LLEAP software by Laerdal Medical™ was utilized for this study. The study population consisted of 20 students from campus 1 and 20 students from campus 2. Students voluntarily signed up for a high-fidelity simulation scenario. There was a mix of second- and third-year students, some of whom had already completed an OB/GYN clerkship and some of whom had not. Students consented to participate in an OB/GYN-related simulation by signing a form that described risks, benefits, COVID-19 precautions, the voluntary nature of their participation, and the confidentiality of the study. The study was vetted by the Institutional Review Board (IRB#: 2020-0076).

### Simulation Scenario

Each participant was assigned a randomized four-digit code to ensure de-identification. Students completed a pre-simulation survey 24 hours prior to the scenario, which included questions about their demographics and asked them to rate their perceived confidence level. The participants were told the simulation would be about PPH, and they were provided with educational materials about PPH, created by the researchers, to guide their preparation. The researchers’ main goal with the simulation was to create a postpartum hemorrhage scenario that could be replicated across two geographically distinct campuses. The primary educational goal for participants in the simulation was to understand the “Four T’s” of PPH: tone, trauma, tissue, and thrombosis.^10^ Secondary educational goals consisted of participants knowing basic treatments for the Four T’s, including usage of potential medications and their dosages.^10^

Video telecommunication was utilized to connect the researchers involved on both campuses for meetings regarding the planning and implementation of the simulations. It was also used during the scenarios to allow faculty to observe participants remotely, in compliance with COVID-19 precautions. An experienced Faculty member acted as the Facilitator at bedside. The case scenario was set up the same day as the scheduled simulation. The SimMom® Laerdal Training simulation model was used in both locations. The postpartum hemorrhage modules were set up according to the Laerdal Training Guide, “Postpartum Hemorrhage Uterus and Postpartum Hemorrhage (PPH) Installation Guide,” which included placental fragments (Figure 1) placed in the uterus and a boggy uterus bag to simulate decreased uterine tone. The manikin’s synthetic abdominal skin was folded back to attach the uterus to the pelvic components. The blood reservoir was filled to capacity at 800 mL, then the feed tube was connected to the outlet on the pelvic bulkhead ^11^.

**Figure 1.**
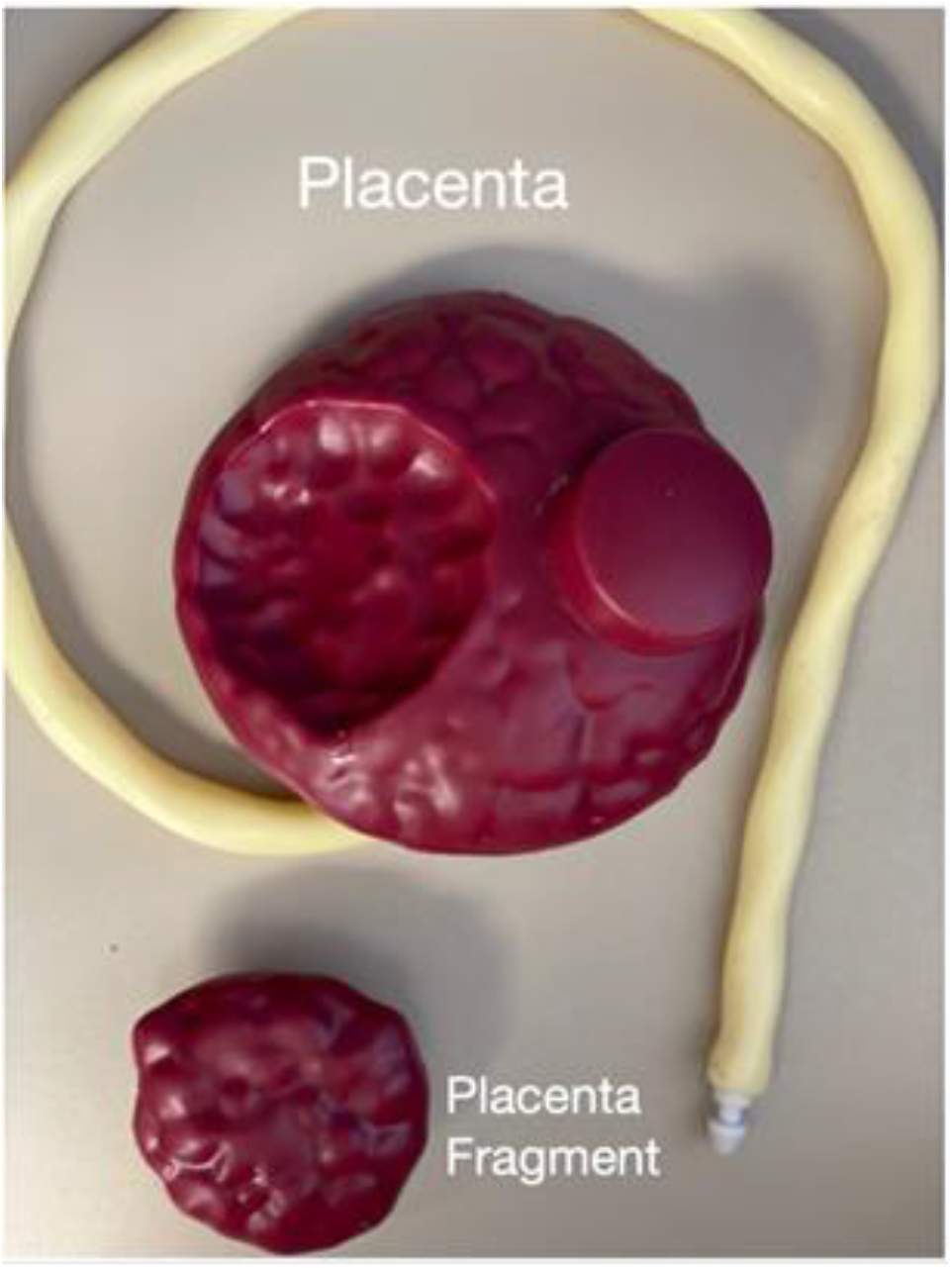
SimMom® placental fragments. Placenta with umbilical cord and placenta fragment. The placenta was placed missing fragment face down in the room to give the students a clue as to the cause of the PPH. The placental fragment was placed inside the SimMom® uterus at the start of the simulation. The placental fragment being removed would mark the successful completion of the simulation scenario.

The rooms were set to resemble a postpartum labor and delivery environment. The manikin was set up supine on a bed, which could be adjusted into a lithotomy position upon student request. Approximately 1000cc of artificial blood was present on and around the under buttocks drapes and in the kick-basin. Speculum, light source, lap sponges, instruments and retractors were available on a Mayo stand and table to the side of the patient. The delivered placenta was placed in a basin facedown with the intention to conceal the missing cotyledon. This was done to encourage the student to actively inspect the placenta for integrity. The retained placental piece was attached to the superior and posterior uterine fundus. A case-scenario ^12^ was specific for PPH due to retained tissue with a simulation that lasted a total of 10 minutes. The timing was initiated after students were notified by delivery nurse, “the patient has already lost 1,000 mL of blood.” “Time A” was marked when students identified and verbalized retained placental tissue as the cause of hemorrhage, and “Time B” was when students extracted the retained placental fragment. Students were observed and subsequently debriefed by facilitators and researchers in the roles described in Table 1.

**Table 1.**
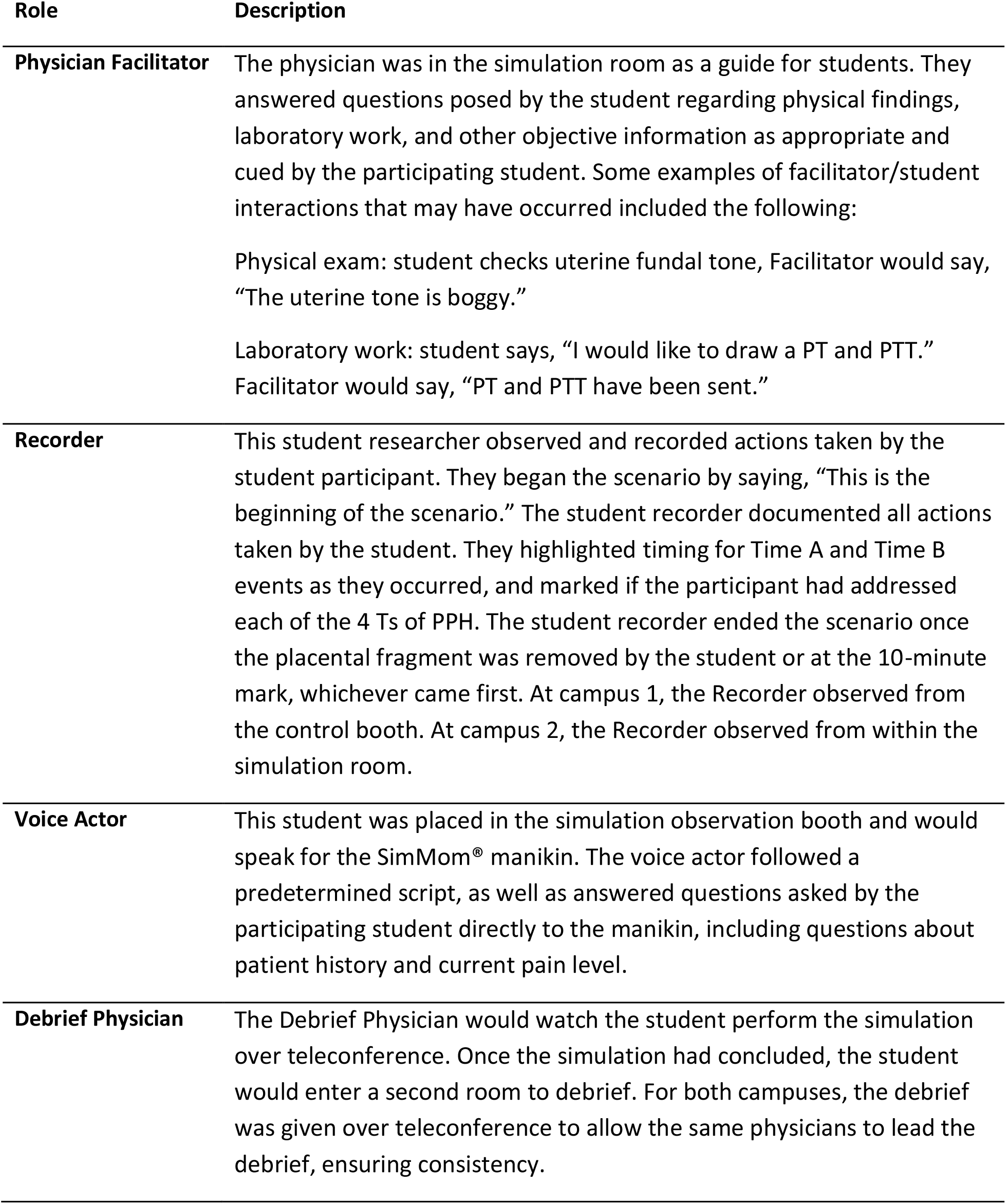
Roles and their descriptions.

Procedural schedule for running PPH simulation allowed for time slots 30 minutes long: 5 minutes for the student to arrive and don personal protective equipment (PPE), 10 minutes per student to perform the simulation, 15 minutes to debrief, and 5 minutes to reset the simulation room for the next student. Staggering two different debrief staff teams allowed for one to view the current simulation as the other debriefed the previous simulation. A graphic schedule is displayed in Figure 2.

**Figure 2.**
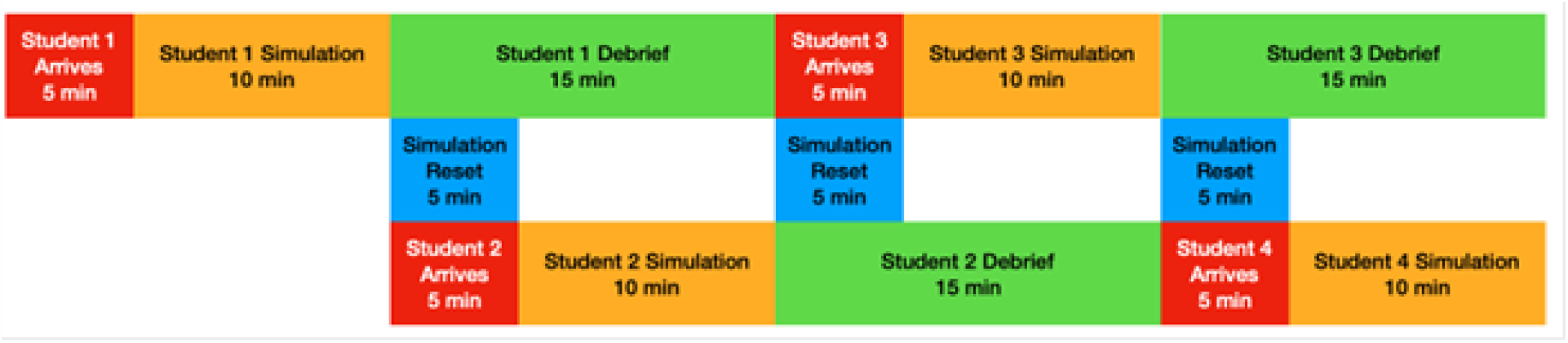
Staggering schedule for debriefing and running the simulation on student teams. This graphical representation demonstrates the scheduling of simulations with the use of two different debrief physicians. Time slots were 30 minutes long: 5 minutes for the student to arrive and don PPE, 10 minutes per student to perform the simulation, 15 minutes to debrief, and 5 minutes to reset the simulation room that occurs when the next student arrives. During the debrief of student one, student two was prepared and ran the simulation. This overlapping schedule was optimal as it decreased the time between the running of each simulation and reduced the time the mannequin was not unuse: overall increasing the number of students able to complete the simulation in the same amount of time.

Due to the experiment being conducted during the COVID-19 pandemic, there was a need to limit the exposure and contact between the researchers, simulation staff and the students. Thus, only one student was performing the simulation at any given time. All involved wore isolation gowns, clean nonsterile gloves, eye protection and a standard surgical mask. Scrub caps and shoe coverings were worn in addition to standard COVID-19 PPE.

### Performance assessment questionnaire

The questionnaire consisted of statements about students’ simulation experience. Students were asked to either agree or disagree with each statement using the following scale: Strongly disagree, Disagree, Neutral, Agree, and Strongly agree. These student answers were correlated to the corresponding numbers for data analysis purposes: 1-Strongly disagree, 2-Disagree, 3-Neutral, 4-Agree, 5-Strongly agree. A copy of the questionnaire is available in Supplementary file 1 and 2.

### Statistical analysis

All data was evaluated using linear models where performance times and overall scores as dependent variables were evaluated for association to demographics, campus, previous experience in obstetrics, and the parameters evaluated by the specific questions. Confidence and comfort perception by the students were analyzed as independent variables. All statistical analyses were performed on SAS/STAT v.9.4 with significant associations declared at a confidence level of P≤0.05 (Table 2).

**Table 2.**
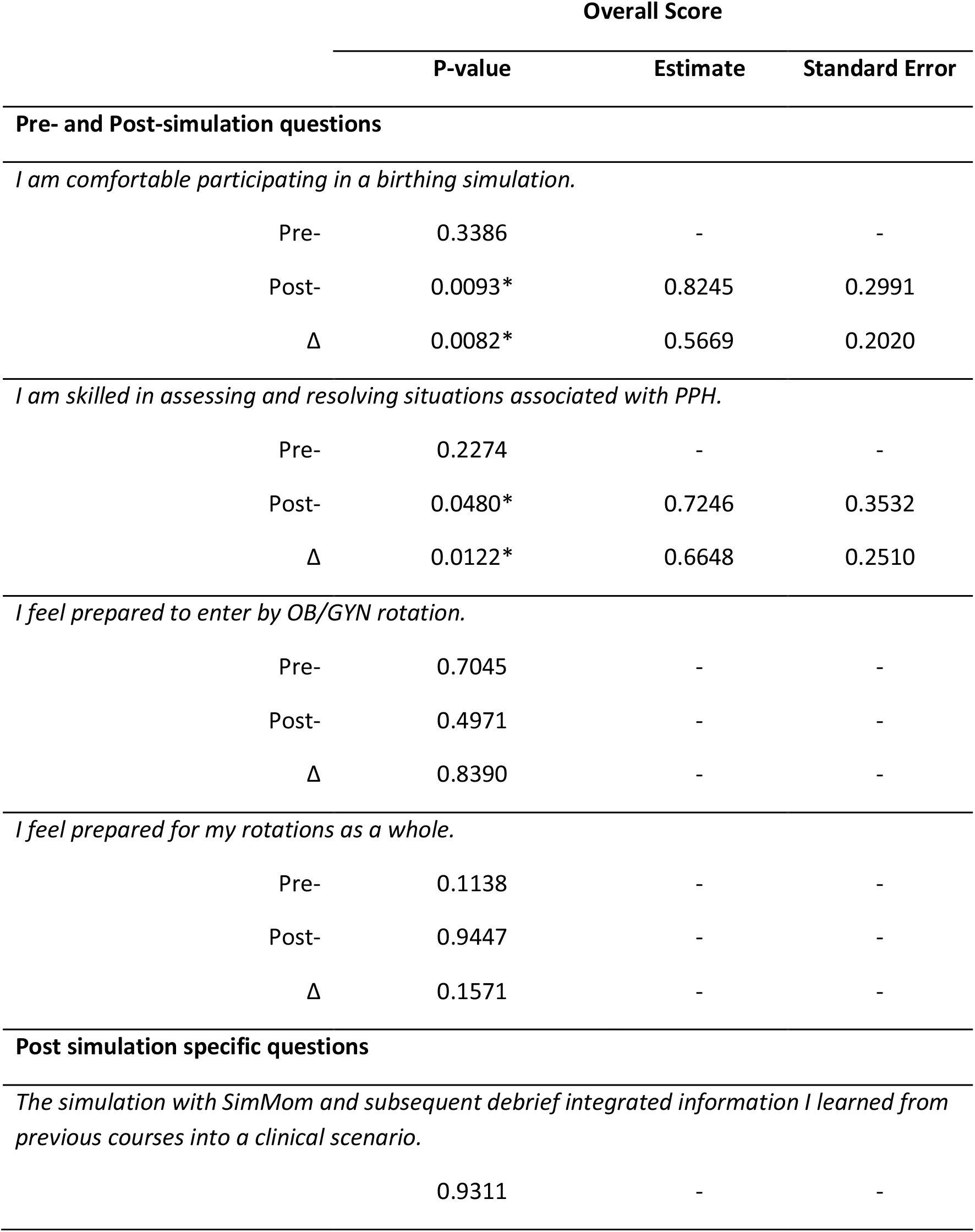
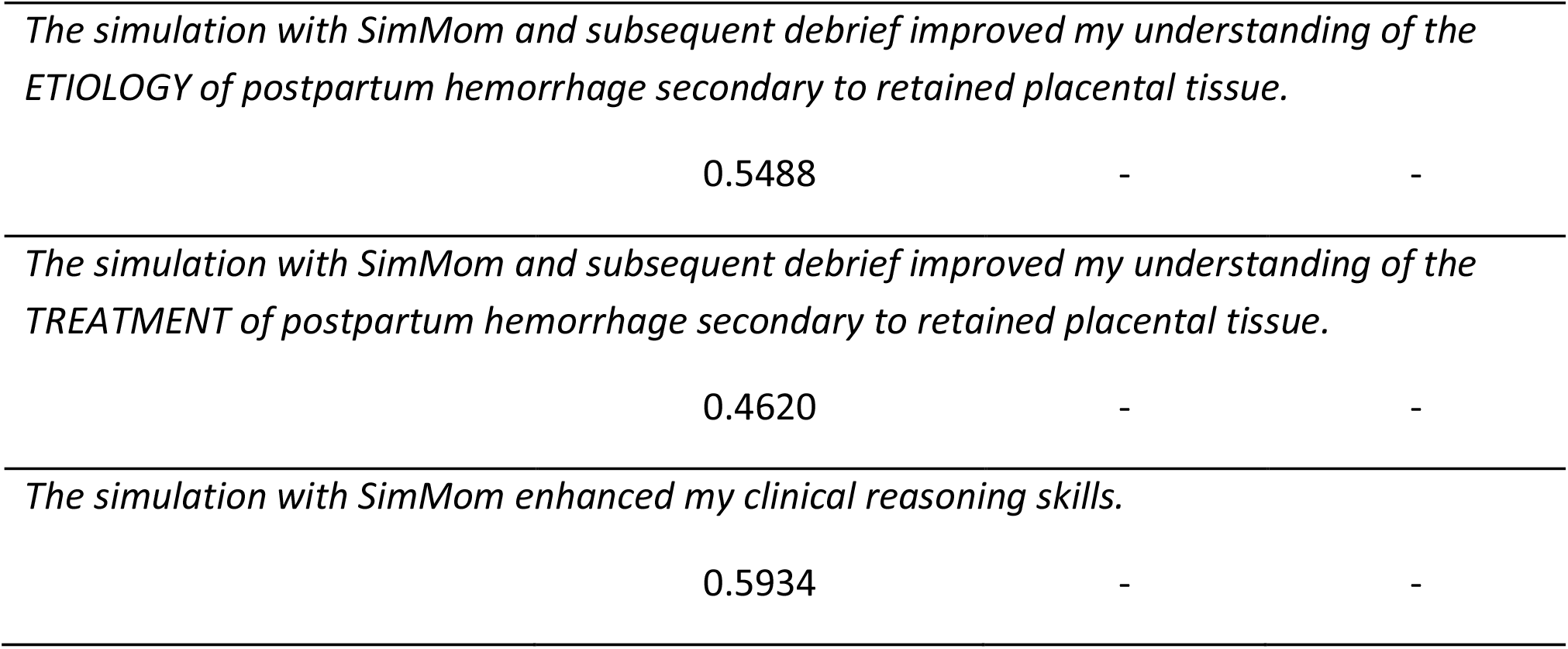
P-values for the students pre and post simulation associations to the overall score for the qualitative evaluation of actions performed during the procedure. Each of the parameters was evaluated independently. Delta values (Δ) evaluates the change from pre- to post-simulation answers. Significance of an association was declared at P≤0.05 and is indicated by an asterisk (*). Only significant estimates and their standard errors are shown.

## RESULTS

Results were derived by comparing the performance of Time A and Time B to demographics, prior simulation, medical, or obstetric experience, demographics, and pre- and post-simulation confidence levels. Each campus had 20 participants with a total n of 40; the age ranges were 26-38 and 23-33 for campus 1 and 2 respectively. On campus 1, 85% of the students were females, 10% had obstetric experience, 45% had completed their OB/GYN rotation, and 45% had experience with a high-fidelity simulation manikin. On campus 2, 50% of the participants were female, and 2% preferred not to answer. 15% percent of the participants had obstetric experience, 35% had experience with a high-fidelity simulation manikin, and 15% had completed their OB/GYN rotation. Despite the efforts to maintain consistent simulations between the two campuses, there was a significant difference between the two campuses for the main time measurements at A and B (P=0.0450 and P=0.0112 respectively).

Although the procedures, scripts, and timings were identical between the two campuses, there was a significant difference discovered in the time recordings. The time it took from the start of the simulation to the time the student evaluated the tone of the uterus average times were 133.444s and 97.45s for campus 1 and campus 2 respectively (P=0.0499). The time it took from the start of the simulation to the time the student recognized retained tissue was the cause of the PPH (Time A) was measured with average times were 233.71s and 375.16s for campus 1 and campus 2 respectively (P= 0.0459). This data has a confounding variable of the 13/20 students in campus 1 and 2/20 students in campus 2 who did not recognize that retained tissue was the cause of the PPH within the 10 min time limit. When changed to binary distribution, 1 representing the completion of the time A mark and 0 representing non-completion of the time A mark, the average was 0.35 and 0.9 for campus 1 and campus 2 respectively (P= 0.0007). The time from the start of the simulation to the time the student removed the retained tissue (Time B) was also measured. Average times were 211.40s and 383.52s for campus 1 and campus 2 respectively (P=0.0186). This data has a confounding variable of the 15/20 students in campus 1 and 3/20 students in campus 2 who did not remove the retained tissue within the 10 min time limit. When changed to binary distribution, 1 representing the completion of the time B mark and 0 representing non-completion of the time B mark, the average was 0.25 and 0.85 for campus 1 and campus 2 respectively (P=0.0009).

Successful completion of the simulation was found to have significance if the student noted previous obstetrical experience in the pre-simulation questionnaire. This affected both the time it took from the start of the simulation to the time the student recognized that retained tissue was the cause of the PPH (Time A) (P=0.0454) as well as the time it took from the start of the simulation to the time the student removed the retained tissue (Time B), (P=0.0283).

The results not only included events occurring during the simulation, but also included the analysis of the pre- and post-surveys. For the post simulation and debrief questionnaire, all questions but one showed no significant difference between the two campuses. When asked on a scale of Poor, Fair, Good, Very Good, or Excellent, “Overall, how would you rate your experience with the postpartum hemorrhage scenario using SimMom?” all 40 participants rated the simulation as Good or Better. Twenty-four out of forty participants rated the simulation Excellent. Of these, 13 rated Excellent from campus 1 and 11 rated Excellent from campus 2. When translated to a 1-5 scale with 1 being Poor and 5 being Excellent, average scores were 4.6 and 4.45 for campus 1 and campus 2 respectively (P= 0.379).

In the evaluation of the students’ simulation experience, when students were asked to rate the following statement, “The simulation with SimMom and subsequent debrief integrated information I learned from previous courses into a clinical scenario,” average scores were 4.65 and 4.55 for campus 1 and campus 2 respectively (P=0.724). When asked, “The simulation with SimMom and subsequent debrief improved my understanding of the etiology of postpartum hemorrhage secondary to retained placental tissue,” average scores were 4.55 and 4.3 for campus 1 and campus 2 respectively (P= 0.397). When asked, “The simulation with SimMom and subsequent debrief improved my understanding of the treatment of postpartum hemorrhage secondary to retained placental tissue,” average scores were 4.6 and 4.75 for campus 1 and campus 2 respectively (P= 0.562). When asked, “The simulation with SimMom enhanced my clinical reasoning skills,” average scores were 4.65 and 4.5 for campus 1 and campus 2 respectively (P= 0.853). When asked, “I am comfortable participating in a birthing simulation,” average scores were 3.8 and 4.44 for campus 1 and campus 2 respectively (P=0.0186). This was the only significant difference between the two campuses on the post simulation and debrief questionnaire. When asked, “I am skilled in assessing and resolving situations associated with PPH,” average scores were non-significant, 3.9 and 3.83 for campus 1 and campus 2 respectively (P=1). When asked, “I feel prepared to enter my OB/GYN rotations,” average scores were 3.65 and 3.77 for campus 1 and campus 2 respectively (P=0.563). When asked, “I feel prepared to enter my rotations as a whole,” on a scale from 1 to 5 and average scores were 3.30 and 3.61 for campus 1 and campus 2 respectively (P=0.217).

Two of the survey questions were found to be significantly associated to their overall performance on the qualitative analysis: “I am comfortable participating in birthing simulation” and “I am skilled in assessing and resolving situations associated with PPH” (P=0.0093 and P=0.0480 respectively, Table 2). This was also observed in the delta values for those questions (P=0.0082 and 0.0122 respectively).

## DISCUSSION

The results highlighted the value of simulations in medical education as well as the importance of standardizing simulation procedures to improve future projects. The statement in the confidence survey, “I am comfortable participating in a birthing simulation” was shown to have significance regarding the frequency at which it was rated (Table 2). This illustrates that the simulations performed across two separate medical campuses were valuable in increasing participants’ feelings of confidence in managing childbirth. However, the differences in the timings from the simulations demonstrate that there is a need to standardize scenarios in future projects.

The difference in Time A and Time B between campus 1 and campus 2 was significant, which may have been due to several factors. There may have been students on campus 2 who were already exposed to a prior PPH simulation and had more baseline knowledge. This could have resulted in a faster completion time, as this prior exposure was not controlled for in the study. Moreover, the facilitators may have performed differently on each campus. The facilitator on campus 2 may have given more guidance to students than the script intended, compared to the facilitator on campus 1. Using the same facilitator to answer students’ questions via video telecommunications in both locations could also improve consistency. However, if the simulations must be carried out at the same time and multiple facilitators are needed, then facilitators should better standardize their script and how they lead the students. An example of this would be whether or not facilitators utilize hints to further guide students to completion of the scenario. This could be addressed in future scenarios by having prepared responses for anticipated common student questions, having separate meetings with facilitators to prepare, or having further training with facilitators.

An additional difference across campuses may have been the physical setup of the room. Instruments available to the participants varied in what was available and how they were arranged on the table. There was also a difference in the number of people that were in the room observing participants on each campus. On campus 2, both the facilitator and the recorder were present in the room. On campus 1, only the facilitator was in the room in person, with a recorder observing participants from outside the room through a tinted window. The debrief physician was observing the scenario remotely using video telecommunication. These differences may have affected participant performance.

Although participants found this to be a valuable experience, more detailed standardization is required to increase the effectiveness of this scenario as an educational tool across geographically separated medical school campuses. To better standardize the simulation experience, the environment, facilitator script, and debriefing model should be well-defined. For example, the delivery room could be setup according to a checklist and predetermined schematic then photographed. The schematic and photographs could be compared between the two campuses before continuing. Once confirmed that the campuses are equivalent, the setup schematic and photographs should be used each time the simulation is reset. In addition to providing consistency, this approach would increase efficiency in set-up and aid in tracking consumables.

Few studies exist in investigating simulated retained placenta as cause of post-partum hemorrhage as a learning modality for medical students. Although there were numerous limitations to this study, it provided foundational groundwork for future research in this area. This study could be expanded to include more participants either in a team setting or working individually. Having a team of students on each scenario could allow for better completion of the simulation as the participants can assist one another. Additionally, this alteration would allow for a larger number of students to complete the simulation in a shorter amount of time thus improving efficiency. Moreover, future investigations could evaluate if simulations in addition to didactic education is as effective or greater compared to the didactic material studied alone in long-term retention of the primary educational goal of the “Four T’s” of PPH: tone, tissue, trauma, and thrombosis. If they are of educational benefit above and beyond conventional in-classroom education, this could help justify the extended amount of time needed to complete simulations.

The challenge of setting two identical obstetric simulation scenarios across two geographically separated medical campuses can be successful but would benefit from specific standardization. With the improvements suggested in the discussion section, this study will encourage further research in post-partum retained placenta simulation scenarios for medical students so that future learners can practice skills and enforce knowledge in a risk-free environment.

## Data Availability

All data produced in the present study are available upon reasonable request to the authors

## ACKNOWLEDGEMENTS

The authors would like to thank Jean Bouquet, D.O., David Ross, D.O., Seth Peacock, M.D Danielle Glaze, D.O., Lauren Galligani, OMS III, Chasity Edwards, SUDC, CHSE, and Tariq Al-Shanteer, CHSOS for their advice and support in the realization of this study.

## Notes

### Competing Interest Statement

The authors have declared no competing interest.

### Funding Statement

This study did not receive any funding

### Author Declarations

Rocky Vista University, Institutional Review Board

### Summary of Updates

First author changed her legal name, the revision only addresses that.

