## Supplementary file 1 for "Standardization of a High-Fidelity Postpartum Hemorrhage Simulation Scenario Across Two Geographically Separated Campuses"

**PRE-SIMULATION DEMOGRAPHICS SURVEY**

1. ID #: _ _ _ _
2. Which is your primary campus?

3. Gender
    a. Female
    b. Male
    c. Non-binary
    d. I prefer to self-describe: _______
    e. I prefer not to answer
4. Age: *rolling list to select exact age*
5. Race: (checkboxes to select more than one)
    a. American Indian or Alaskan Native
    b. Asian
    c. Black/African-American
    d. Native Hawaiian or Other Pacific Islander
    e. White
    f. I prefer not to answer
6. Ethnicity:
    a. Hispanic or Latino
    b. Not Hispanic or Latino
    c. I prefer not to answer

*7. Before starting medical school, did you have any experience in healthcare and medicine? (ex. Shadowing, scribing, working as a medical assistant, etc)*
    *a. No, I do not have any background experience in healthcare.*
    *b. Yes; please briefly describe: ________*

*8. Before starting medical school, did you have any experience in obstetrics? (ex. Midwife or doula, etc)*
    *a. No, I do not have any background experience in obstetrics.*
    *b. Yes; please briefly describe: ________*

*9. Have you ever participated in a high-fidelity simulation with a manikin?*
    *a. No*
    *b. Yes; please briefly describe: _______*

*10. This simulation will involve artificial blood. Are you still willing to participate?*
    *a. Yes*
    *b. No*

**PRE-SIMULATION CONFIDENCE SURVEY**

**Rate on a scale of 1-5**

1 - Strongly disagree
2 - Disagree
3 - Neutral
4 - Agree
5 - Strongly Disagree

1.) I am comfortable participating in a birthing simulation.
2.) I am skilled in assessing and resolving situations associated with PPH.
3.) I feel prepared to enter my OB/GYN rotations.
4.) I feel prepared for my rotations as a whole.
