## Supplementary file 2 for "Standardization of a High-Fidelity Postpartum Hemorrhage Simulation Scenario Across Two Geographically Separated Campuses"

**POST-SIMULATION EXPERIENCE SURVEY**

1. ID #: _ _ _ _
2. Overall, how would you rate your experience with the postpartum hemorrhage scenario using SimMom® on X/X/XX?
    a. Excellent
    b. Very good
    c. Good
    d. Fair
    e. Poor
3. How did you prepare for the simulation? ______
4. What did you like about the simulation? _______
5. What did you dislike about the simulation? _______
6. The simulation with SimMom® and subsequent debrief integrated information I learned from previous courses into a clinical scenario.
    a. Strongly agree
    b. Agree
    c. Neither agree nor disagree
    d. Disagree
    e. Strongly disagree
7. The simulation with SimMom® and subsequent debrief improved my understanding of the **etiology** of postpartum hemorrhage secondary to retained placental tissue.
    a. Strongly agree
    b. Agree
    c. Neither agree nor disagree
    d. Disagree
    e. Strongly disagree
8. The simulation with SimMom® and subsequent debrief improved my understanding of the **treatment** of postpartum hemorrhage secondary to retained placental tissue.
    a. Strongly agree
    b. Agree
    c. Neither agree nor disagree
    d. Disagree
    e. Strongly disagree
9. The simulation with SimMom® enhanced my clinical reasoning skills.
    a. Strongly agree
    b. Agree
    c. Neither agree nor disagree
    d. Disagree
    e. Strongly disagree
10. Do you have any additional comments or feedback? ______

**POST-SIMULATION CONFIDENCE SURVEY**

**Rate on a scale of 1-5**

1 - Strongly disagree
2 - Disagree
3 - Neutral
4 - Agree
5 - Strongly Disagree

1.) I am comfortable participating in a birthing simulation.
2.) I am skilled in assessing and resolving situations associated with PPH.
3.) I feel prepared to enter my OB/GYN rotations.
4.) I feel prepared for my rotations as a whole.
